# Associations between mosaic loss and schizophrenia or bipolar disorder of young onset

**DOI:** 10.1101/2025.06.13.25329510

**Authors:** Shunsuke Uchiyama, Takeo Saito, Xiaoxi Liu, Yuki Ishikawa, Keiko Hikino, Masashi Ikeda, Giulio Genovese, Nakao Iwata, Chikashi Terao

**Affiliations:** Laboratory for Statistical and Translational Genetics, RIKEN Center for Integrative Medical Sciences, Yokohama, Japan; Department of Allergy and Rheumatology, Nippon Medical School Graduate School of Medicine, Tokyo, Japan; Department of Psychiatry, Fujita Health University School of Medicine, 1-98 Dengakugakubo, Katsukake-cho, Toyoake, Aichi, 470-1191, Japan; Laboratory for Pharmacogenomics, RIKEN Center for Integrative Medical Sciences, Yokohama, Japan; Department of Psychiatry, Nagoya University Graduate School of Medicine, 65 Tsurumai-cho, Showa-ku, Nagoya, Aichi, 466-8560, Japan; Program in Medical and Population Genetics, Broad Institute of MIT and Harvard, Cambridge, MA, USA; Department of Genetics, Harvard Medical School, Boston, MA, USA; Clinical Research Center, Shizuoka General Hospital, Shizuoka, Japan; Department of Applied Genetics, The School of Pharmaceutical Sciences, University of Shizuoka, 52-1 Yada, Suruga Ward, Shizuoka, 422-8526, Japan

**Author notes:** These authors contributed equally: Shunsuke Uchiyama, Takeo Saito. Correspondence author: Chikashi Terao MD, PhD Laboratory for Statistical and Translational Genetics, Center for Integrative Medical Sciences, Riken, Yokohama 230-0045, Japan, Nakao Iwata MD, PhD Department of Psychiatry, Fujita Health University School of Medicine, 1-98 Dengakugakubo, Katsukake-cho, Toyoake, Aichi, 470-1191, Japan.

## Abstract

Mosaic chromosomal alterations (mCAs) accumulate in the brain tissues and are associated with psychiatric disorders. The association between mCAs in circulating blood and schizophrenia (SCZ) and bipolar disorders (BD) has not been fully evaluated. We detected mCAs from blood samples in 2,470 SCZ, 3,732 BD, and 177,773 control subjects. The associations between mCAs and SCZ or BD were evaluated using age-adjusted logistic regression models, further evaluated in age subgroups. The associations were further evaluated based on age at onset. We analyzed the associations between high cell fraction (CF) mosaic events (CF-mCAs >5% or CF-mCAs >10%) and SCZ or BD in the same way. Furthermore, we assessed the interaction between mCAs and genetic risk scores for SCZ or BD. Autosomal mCAs, especially mosaic loss events, increased in both SCZ and BD (SCZ; OR=1.78, P=4.9×10^-6^, BD; OR=1.41, P=0.0025). These associations were highlighted in the young-age subgroup (SCZ; OR=7.01, P=1.7×10^-16^, BD; OR=4.01, P=2.9×10^-8^). We observed the strong effect sizes for association with the diseases at young onset (SCZ; OR=2.51, P=5.0×10^-5^). In addition, the effect sizes of losses increased in a CF-dependent manner in both SCZ and BD. Loss events with high cell fraction interacted with polygenic risk score in SCZ (P=0.0098). SCZ or BD were characterized by the presence of a high burden of mosaic losses in blood, especially in onset of young age, suggesting the common somatic pathophysiological mechanisms between these psychiatric diseases. The possible interaction between losses and PRS for SCZ supports the genetic and environmental cross-talk in SCZ.

## INTRODUCTION

Schizophrenia (SCZ) and bipolar disorder (BD) are complex traits in which various causes and mechanisms, including genetic and environmental components, contribute to the onset [1]. Recent studies have identified nearly hundreds of susceptibility loci associated with SCZ and BD [2]. From the viewpoint of environmental factors, many factors such as obstetric complications or infections, have been investigated so far [1]. However, these components can explain a part of phenotypic variance and majority of causes are still underexplored.

Somatic mutation is a part of yet-to-be-identified causes of psychiatric diseases. Historically, due to easy access to samples, somatic mutations have been investigated in blood cells in the context of haematological malignancy and age-related diseases [3]. However, recent studies have found that somatic mutations also occur in non-blood cells, including neurons in the normal human brains through aging, contributing to neural diversity [4]. Bae and Lodato et al. found that somatic mutations occur in neural progenitor cells in the brain during the pregastrulation period [5] and accumulate by age [6]. Additionally, previous studies have found that somatic mutations are associated with neurodevelopmental diseases [7]. Among the somatic mutations, mosaic chromosomal alterations (mCA) are known to be the clonal cellular expansion with somatic mutations, which range to the wide chromosomal rearrangements [8]. Particularly, mCAs, which occur in the brain at an early developmental period, are reported to be associated with various neurodevelopmental diseases including SCZ [9, 10]. Specifically, recent reports have identified that mCAs in circulating blood, particularly somatic losses, are associated with SCZ in European and Chinese populations, possibly by altering the gene function [9, 10]. For example, recurrent somatic deletions of NRXN1, which encodes a presynaptic adhesion protein, were observed in patients with SCZ [9]. In addition to the blood samples, Maury et al. have found that somatic mosaicism is enriched in open chromatin regions in SCZ brains [11].

Previous studies [9, 10] have analyzed the association between mCAs in circulating blood and psychiatric diseases due to easy access to samples. However, there are challenges regarding the interpretations of the results. The first challenge is that we can only capture mCAs with high cell fraction (CF) due to technical limitations using intensity data of DNA microarray, in contrast to deep depth sequencing data. Given that high CF of mCAs in blood were shared in brain cells and other cells [12], studying mCAs in circulating blood would reflect mCAs in the brain to some extent. Another challenge is that it is necessary to remove age-related mCA events when we focus on the disease-related mCAs. In the previous reports [9, 10], the authors could not definitively compare frequencies of mCAs by adjusting age of samples in their statistical model to remove age-related somatic mutations since many of the samples analyzed in the previous study lacked information on age (alternatively, the authors filtered mCAs that are associated with clonal haematopoiesis of indeterminate potential (CHIP)). In addition, the interactions between genetic and somatic architecture have never been explored.

Here, we analyzed the associations of mCAs (autosome, mosaic loss of X (mLOX), and mosaic loss of Y (mLOY)) with SCZ and BD derived from two datasets using logistic regression models adjusted by covariates including age. We also evaluated the associations between mCAs and SCZ or BD subdivided based on the onset age. Our findings suggest that mCAs substantially increase in SCZ and BD of young onset, showing the possible interaction between mosaic losses and genetic risk with SCZ.

## SUBJECTS AND METHODS

### Study Participants

In this study, we recruited 2,555 patients with SCZ and 3,998 patients with BD from hospitals in Japan [13–15] (Fig. 1). The control samples were recruited from the BioBank Japan cohort (BBJ cohort) [16]. The psychiatric disorders were not included in the 47 targeted diseases in the BBJ cohort [16]. The details of the definitions of cases have been described in our previous papers [13–15]. Two experienced psychiatric doctors diagnosed SCZ or BD according to the DSM-IV and DSM-IV-TR criteria, respectively, through the unconstructed medical interviews with patients and their families and a review of the medical records. We removed the cases who have any one of the following criteria: cases with other physical problems, mood disorders, substance abuse, neurodevelopmental disorders, epilepsy, or known mental retardation. All patients provided informed consent, and this study was approved by the local ethics committee of each institute. Based on the previous reports [17], we defined SCZ with the age of the disease onset ≧45 or <45 years as late onset SCZ or early onset SCZ, respectively. We also defined BD with the age of the disease onset ≧50 or <50 years as late onset BD or early onset BD in the same way [18].

**Figure 1.**
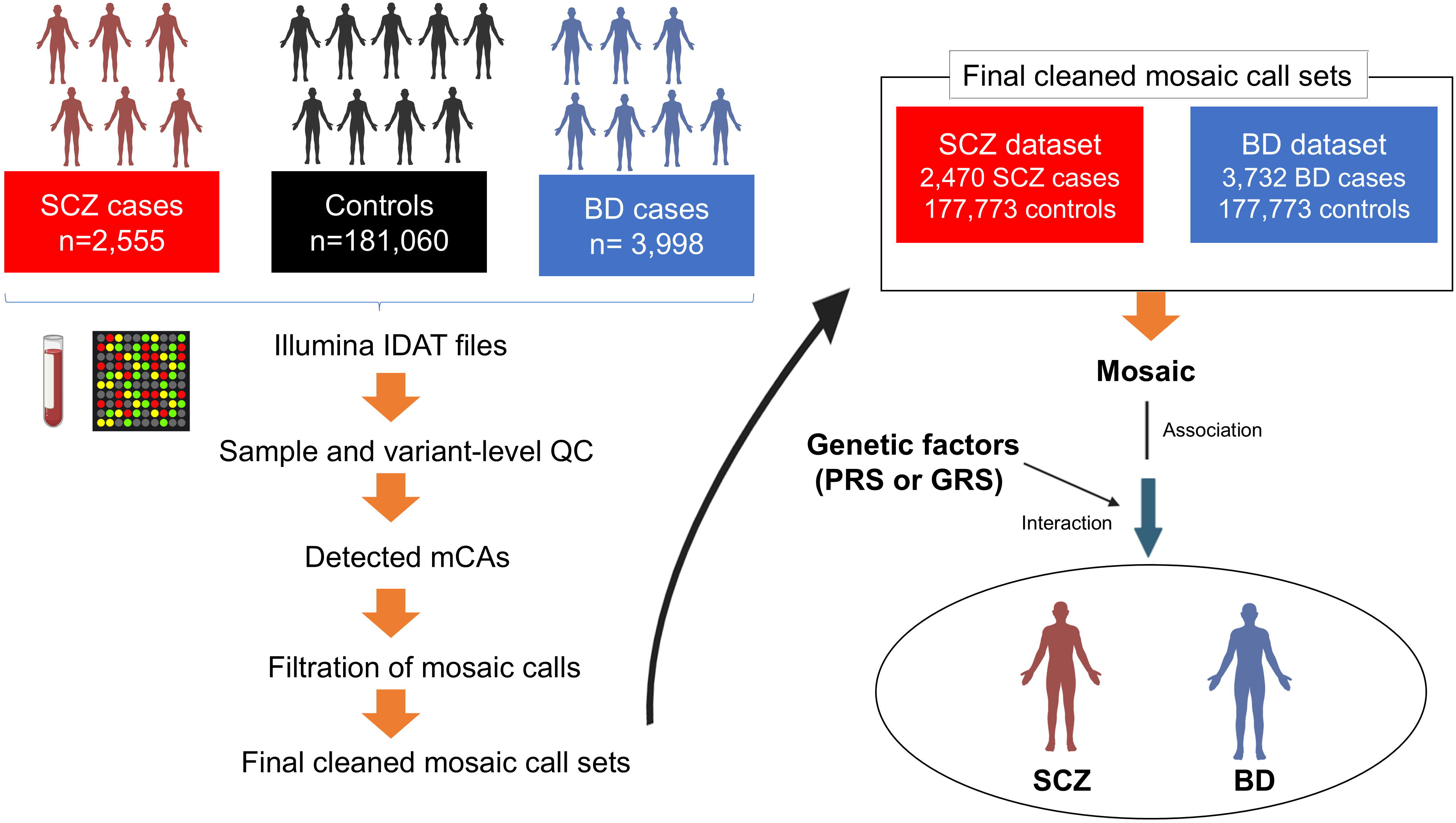
Schematic view of mosaic calling and study design. Method of mosaic calling by the MoChA (left). Study subjects in each dataset (SCZ and BD datasets) (right). SCZ, schizophrenia; BD, bipolar disorder; PRS, polygenic risk score; GRS, genetic risk score.

### Genotyping

We performed the genotyping using the blood DNA of all individuals. The samples of SCZ and BD were genotyped using Human OmniExpress Exome v1.0 (OEEv1.0), OEEv1.2, OEEv1.4, or OEEv1.6. Details of genotyping of the BBJ samples have been described elsewhere [19]. Briefly, BBJ samples were genotyped using OEEv1.0, OEEv1.2, or a combination of Illumina HumanOmniExpress v1.0 and HumanExome v1.0, 1.1 (OE+HE). The variants on the HumanExome were not used in detection of mCAs and included to construct polygenic risk score (PRS) (see below).

### Variant- and Sample-level quality controls and detection of mosaic events

At first, we performed the variant and sample-level QC according to the methods in our previous studies (Fig. 1) [20]. For the variant-level QCs, we removed the variants that met any of the following criteria: variants with variant call rate < 0.97, variants with excess heterozygosity (p<1e-6, Hardy-Weinberg equilibrium test), variants falling in segmental duplications with low divergence (<2 %) and variants with allele frequency difference from those of the reference panel >3.0 %. For a imputation panel, we used a previously constructed imputation panel from the phase 3 1000 genome project ver.5 (1KGP3ver5) (http://www.internationalgenome.org/data) [21] combined with high-depth WGS data from 3265 Japanese subjects from BBJ (J3K), which enables rare variant detection [22]. For the sample-level QCs, we removed samples of the outliers from East Asian populations determined by the principal component analysis and genetically identical samples (PIHAT > 0.9 estimated by PLINK 1.9) [23]. We also removed the samples with sample call rate < 0.97 or samples with phased BAF autocorrelation > 0.03. After the sample and variant-level QCs, we performed phasing using SHAPEIT4 using all samples together across overlapping genome windows [24]. After the phasing, we detected mCAs by the long-range haplotype phasing method using MoChA WDL pipelines. After the MoChA, we excluded samples who met one of the following criteria: samples less than 18 years old at genotyping, samples with genotype and phenotype sex discordance, and samples with missing data on their sex and/or age status. We also removed samples with a past medical history of haematological malignancies only in the BBJ samples because we observed a strong association between autosomal mCAs and haematological malignancy [19]. The filtering of mosaic calls was set as follows, and mosaic chromosomal alterations that have met one of these criteria were removed: germline copy number polymorphism (CNPs), constitutional mosaic events (lod_baf_phase <10), and germline duplications (length < 500 kbp and relative coverage > 2.5). To filter out XXY and XXX samples, we restricted mLOX and mLOY with estimated ploidy less than 2.5. Finally, we identified the autosomal mCAs (subdivided by loss, copy-neutral loss of heterozygosity (CN-LOH), or gain according to the copy number state) and mCAs on sex chromosomes (mLOY in males and mLOX in females). We also detected the undetermined mCAs, which we could not determine the copy number state, and they were categorized in the presence of autosomal mCAs. For each loss, CN-LOH, and gain events that passed mosaic-level QCs, we calculated the cell fraction (CF) of these mosaic events based on these formula (loss: 4*bdev/(1+2*bdev), CN-LOH: 2*bdev, gain: 4*bdev/(1-2*bdev), where bdev represents the BAF deviation estimated from 0.5). The 2022-12-21 version of the MoChA WDL pipeline was used to detect autosomal mCAs, mLOX, and mLOY. Finally, we made cleaned mosaic call sets and the present study consists of the SCZ datasets (2,470 patients with SCZ and 177,773 controls) and the BD datasets (3,732 patients with BD and 177,773 controls) (Fig. 1).

### Construction of polygenic risk score for SCZ and genetic risk score for BD

We calculated the PRS of SCZ using PRS-CS [25]. As the inputs of PRS-CS, we used the previously published summary statistics of the SCZ-GWAS [26] and LD reference panel of East Asian clusters constructed using the 1000 Genomes project phase 3 samples. At first, we inferred the posterior effect sizes, and we extracted the effect sizes of the SNPs shared among the present SCZ datasets and the previous summary statistics [26]. Then, the posterior effect sizes of the risk allele were multiplied by the corresponding genotype dosages of the risk alleles and summed for each individual. Regarding the samples with BD, we calculated genetic risk score (GRS) since we could not use the available public summary statistics of BD GWAS in East Asian clusters with large sample sizes. For the calculation of GRS, we first retrieved the effect sizes of 64 lead SNPs from previously published BD GWAS in EUR populations [27]. Only 58 lead SNPs shared with the present BD datasets were used. Then, we multiplied the effect sizes by the individual genotypes of the risk allele and summed for each individual. The imputed genotypes were used for the calculation of the genetic risk scores for SCZ and BD. For SNP imputation, we excluded the variants which have met one of any following criteria: variants with call rate < 0.97, variants with allele frequency difference between the imputation panel > 3.0%, variants with hardy-Weinberg equilibrium p-value (HWE-P) < 1.0×10^-6^, or variants with minor allele frequency (MAF) < 0.005. We used the phase 3 1000 genome project ver.5 (1KGP3ver5) (http://www.internationalgenome.org/data) [21] combined with high-depth WGS data from 3265 Japanese subjects from BBJ (J3K) as a imputation panel. After the imputation, we removed the variants with low imputation accuracy (R^2^ < 0.3). The only samples who have passed the sample-level QCs (as shown above) were used to calculate PRS and GRS.

### Statistical analysis

To evaluate the associations between mCAs and SCZ or BD, we performed the logistic regression analysis as follows:

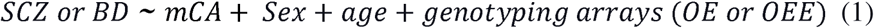

We applied this model separately to the autosomal mCAs, loss, gain, CN-LOHs, mLOX, and mLOY. Only males and females were used in mLOX and mLOY analysis, respectively without sex as a covariate. We further stratified samples in both datasets into four subgroups by age (< 40 ages, 40-49 ages, 50-59 ages, and > 60 ages), then analyzed the associations between mCAs and SCZ or BD in each subgroup in the same manner as described above. To evaluate whether these associations differed between early- and late-onset cases, we subdivided SCZ and BD into young onset or late onset phenotypes respectively, according to the definitions determined in the Methods. In this analysis, we only used samples with information of the onset age of these diseases. Further, we analyzed the associations between autosomal mCA-cell fraction (autosomal mCA-CF) > 5 or 10 % and SCZ or BD using the same logistic regression models as the equation (1). The association between loss-cell fraction (loss-CF) > 5 or 10% and SCZ or BD were also assessed. These associations with high CF of mCAs were also evaluated based on age at onset. We also evaluated the frequency of gene involvement of mosaic loss events in patients with SCZ or BD by the Cochran-Mantel-Hanzel (CMH) test adjusted by sex and age using the complete gene list (glist-hg38) (https://www.cog genomics.org/plink/1.9/resources) (analyzed gene number= 25,009) as the reference and further evaluated based on age at onset.

Next, we analyzed the association between the SCZ or BD risk factors and these disease phenotypes using logistic regression models as follows:

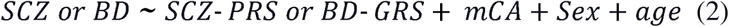

We used the genetic factors (SCZ-PRS or BD-GRS) and mCA as the risk factors in this formula. Autosomal mCA and mosaic loss events were applied separately in this formula. To avoid overfitting, we only used samples with PI_HAT≦0.25 by identity-by-descent (IBD) detection. We also assessed the association between autosomal mCA or losses and PRS or GRS, adjusting for sex, age, and age^2^ to ensure no associations between them.

Finally, to evaluate the effect of interaction between mCA and PRS on SCZ, we stratified the SCZ cases and controls into six subgroups based on the PRS and mosaic status. We used low-PRS (≦25%) and not low PRS status (>25%) according to the distributions of PRS among SCZ cases. The mCA status was subdivided into mCAs negative, mCAs positive, and mCAs-CF > 5%. After categorizing these subgroups, we assessed the risks of SCZ in each subgroup, setting the low-PRS and mCA negative subgroup as a reference by the logistic regression models adjusted sex and age as follows:

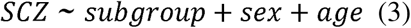

To further evaluate the possible interaction between mCA and the genetic risk scores (PRS for SCZ and GRS for BD), we constructed the following two logistic regression models (one without interaction terms and the other with interaction terms) and evaluated the homogeneity of effects of mCA on SCZ between low PRS and not low PRS group by one-way ANOVA:

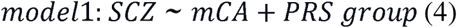

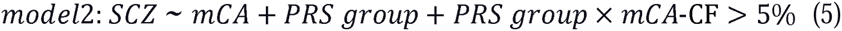

where SCZ (0: non-SCZ, 1: SCZ), PRS group (0: low, 1: high), and mCA-CF >5% (0: mCA-negative or mCA-CF <5%, 1: mCA-CF >5%) are all binary traits; hence, is an interaction term (PRS group × mCA-CF >5 %) in Equation (5). In the interactive analysis shown above, we applied to autosomal mCA and loss events separately. In addition to the patients with SCZ, we also evaluated the patients with BD in the same manner, using GRS as the genetic risk.

We set control subjects as a reference in the analyses to evaluate the associations between mCAs or mCA-CF >5 or 10% with SCZ or BD, and the associations between autosomal mCAs or losses and genetic factors for SCZ or BD. The statistical significance of the associations between mCAs or mCA-CF >5 or 10% and SCZ or BD was based on the Bonferroni correction (P<0.05/6 = 0.008). We also used the Bonferroni correction (P<0.05/24 = 0.002) to determine the statistical significance of the associations between mCAs and SCZ or BD in each age subgroup. The statistical significance of not only the associations between mosaic losses and specific locus but also the interactive associations between mCA and genetic risk among the five subgroups was also based on Bonferroni correction (P<0.05/25,009= 1.9×10^-6^, P<0.05/5 = 0.010 respectively). In other logistic regression models, we set the significance threshold at a two-sided P-value of <0.05. All statistical analyses were conducted using the R software (version 3.3.3).

## RESULTS

### Characteristics of study participants

The demographic features of the study participants are shown in Supplementary Table S1. Patients with SCZ or BD were younger than controls in both datasets. The prevalence of females was slightly higher in cases than in controls in both diseases (47.5 % of SCZ and 46.4 % of controls, 51.9 % of BD and 46.4 % of controls, respectively).

### Significant positive associations of mosaic loss events with SCZ or BD

Consistent with the previous findings, we observed any types of mCAs increased by age, regardless of disease status (Fig. 2). Autosomal mCAs, defined by CN-LOHs, loss, gain, and unclassified mosaics in autosomes, significantly increased in SCZ (OR=1.57, P=6.1×10^-11^), adjusted for the covariates including age in the logistic regression model (METHODS) (Table 1 and Fig. 2). Autosomal mosaic loss events significantly drove the associations between autosomal mCAs and SCZ (OR=1.78, P=4.9×10^-6^) (Table 1 and Fig. 2). We did not find a significant association between CN-LOHs or gain and SCZ (Table 1 and Fig. 2). These results were in line with the findings in the previous studies of SCZ [9, 10], while the previous studies lacked the age of samples in their data sets. Of note, we found a similar positive association between BD and autosomal mCAs or mosaic loss events (autosomal mCAs; OR=1.14, P=0.036, mosaic loss; OR=1.41, P=0.0025), while the effect sizes were lower than those of SCZ (Table 1 and Fig. 2).

**Figure 2.**
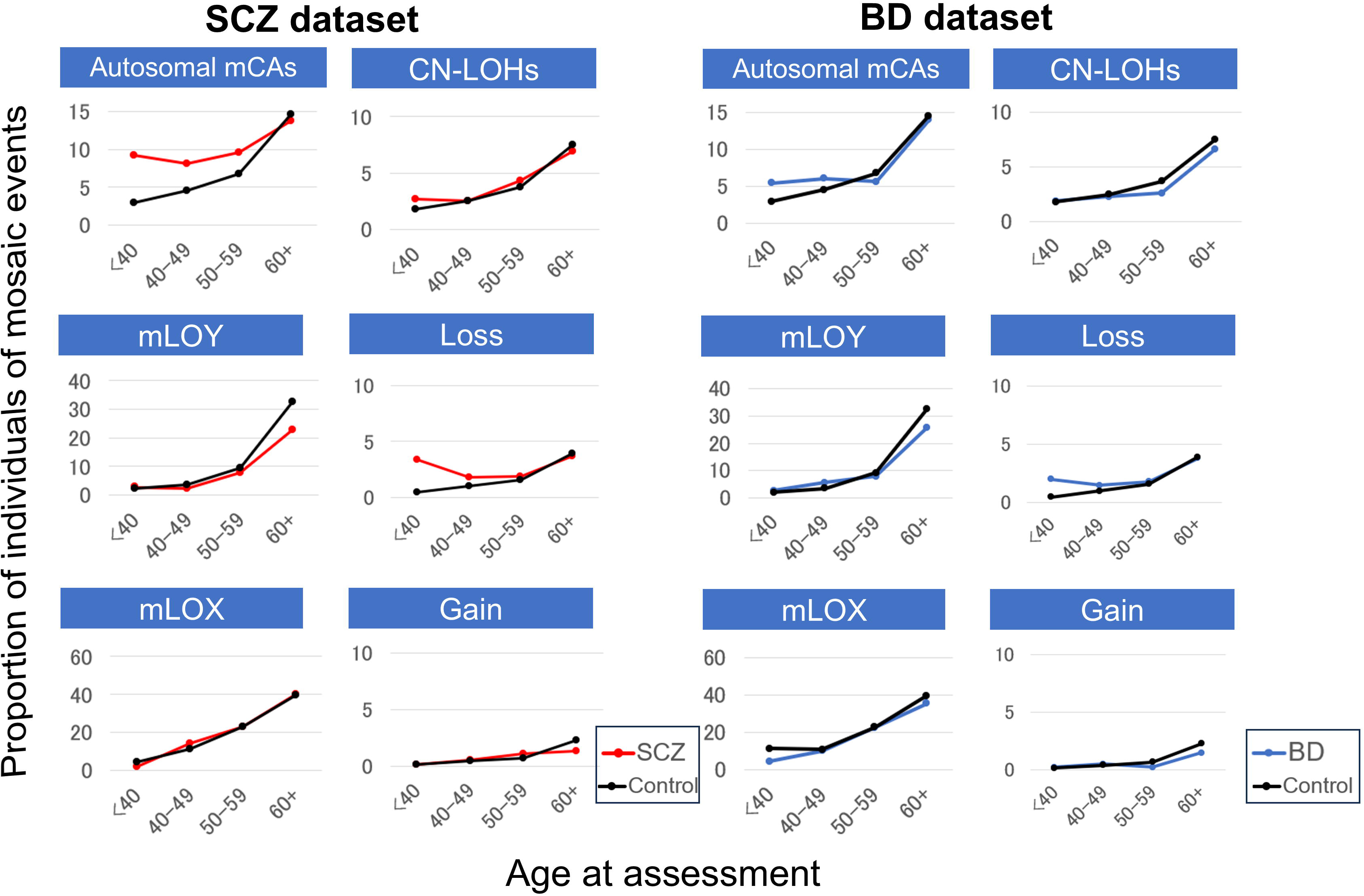
Frequency of different detectable mosaic events stratified by age. The frequencies (means) are indicated for the SCZ dataset (2,470 SCZ and 177,773 controls) and the BD dataset (3,732 BD and 177,773 controls). mCAs, mosaic chromosomal alterations; CN-LOHs, copy-neutral loss of heterozygosity; mLOX, mosaic loss of chromosome X; mLOY, mosaic loss of chromosome Y.

**Table 1.** The associations between different types of mCAs and SCZ or BD in each dataset corresponding to Figure 2.

| SCZ | Cases with<br>mCAs | Controls with<br>mCAs | OR<br>(95 % CI) | P | BD | Cases with<br>mCAs | Controls with<br>mCAs | OR<br>(95 % CI) | P |
| --- | --- | --- | --- | --- | --- | --- | --- | --- | --- |
| Autosomal<br>mCAs | 251/2,470<br>(10.2%) | 20,232/177,773<br>(11.4%) | 1.57<br>(1.37-1.80) | **6.1×10<sup>-11</sup>** | Autosomal<br>mCAs | 288/3,732<br>(7.7%) | 20,232/177,773<br>(11.4%) | 1.14<br>(1.01-1.29) | 0.036 |
| CN-LOH | 99/2,470<br>(4.0%) | 10,518/177,773<br>(5.9%) | 1.11<br>(0.90-1.36) | 0.32 | CN-LOH | 123/3,732<br>(3.3%) | 10,518/177,773<br>(5.9%) | 0.89<br>(0.74-1.07) | 0.23 |
| Loss | 70/2,470<br>(2.8%) | 5,280/177,773<br>(3.0%) | 1.78<br>(1.38-2.26) | **4.9×10<sup>-6</sup>** | Loss | 85/3,732<br>(2.3%) | 5,280/177,773<br>(3.0%) | 1.41<br>(1.13-1.75) | **0.0025** |
| Gain | 17/2,470<br>(0.69%) | 2,967/177,773<br>(1.7%) | 0.86<br>(0.53-1.39) | 0.54 | Gain | 23/3,732<br>(0.62%) | 2,967/177,773<br>(1.7%) | 0.75<br>(0.50-1.14) | 0.18 |
| Female mLOX | 221/1,174<br>(18.8%) | 25,277/82,461<br>(30.7%) | 0.99<br>(0.85-1.17) | 0.95 | Female mLOX | 330/1,936<br>(17.0%) | 25,277/82,461<br>(30.7%) | 0.90<br>(0.79-1.02) | 0.11 |
| Male mLOY | 105/1,296<br>(8.1%) | 22,829/95,312<br>(24.0%) | 0.74<br>(0.60-0.91) | **0.0047** | Male mLOY | 178/1,796<br>(9.9%) | 22,829/95,312<br>(24.0%) | 0.83<br>(0.70-0.98) | 0.025 |

OR, odds ratio referring to non-psychiatric disease controls; P value in bold indicate statistical significance based on Bonferroni correction (P<0.008). SCZ, schizophrenia; BD, bipolar disorder; mCAs, mosaic chromosomal alterations; CN-LOH, copy number-loss of heterozygosity; mLOX, mosaic loss of chromosome X; mLOY, mosaic loss of chromosome Y.

### Significant negative associations of mLOY with SCZ

Regarding the mosaic events on sex chromosomes, mLOX was not associated with SCZ or BD. On the other hand, mLOY showed a significant negative association with SCZ (OR=0.74, P=0.0047). We also observed a negative association between mLOY and BD, while they did not meet the Bonferroni corrected significance (OR=0.83, P=0.025) (Table 1 and Fig. 2).

### Highlighted positive associations between autosomal mCAs or loss with SCZ or BD in the young-age subgroup

Since the prevalence of autosomal mosaicism in SCZ showed notable differences between cases and controls in a young-age subgroups (Fig. 2), we assessed the associations between mosaics and SCZ in each of age subgroups (putting age as a covariate in spite of the narrow range of age differences in each subgroup). As a result, the associations between SCZ and autosomal mCAs or mosaic losses were highlighted in the young-age subgroup and decreased depending on age (Fig. 3 left). In contrast, the associations between SCZ and the other mosaic events (CN-LOHs, gain, mLOX, and mLOY) did not show an age-dependent decreasing trend (Supplementary Fig. S1). Likewise, the association between BD and autosomal mCAs or losses steadily decreased in the same manner (Fig. 3 right and S2). Taken together, these results indicate that autosomal mCA, particularly mosaic losses, increased in SCZ or BD, and these associations were highlighted in the young-age subgroup.

**Figure 3.**
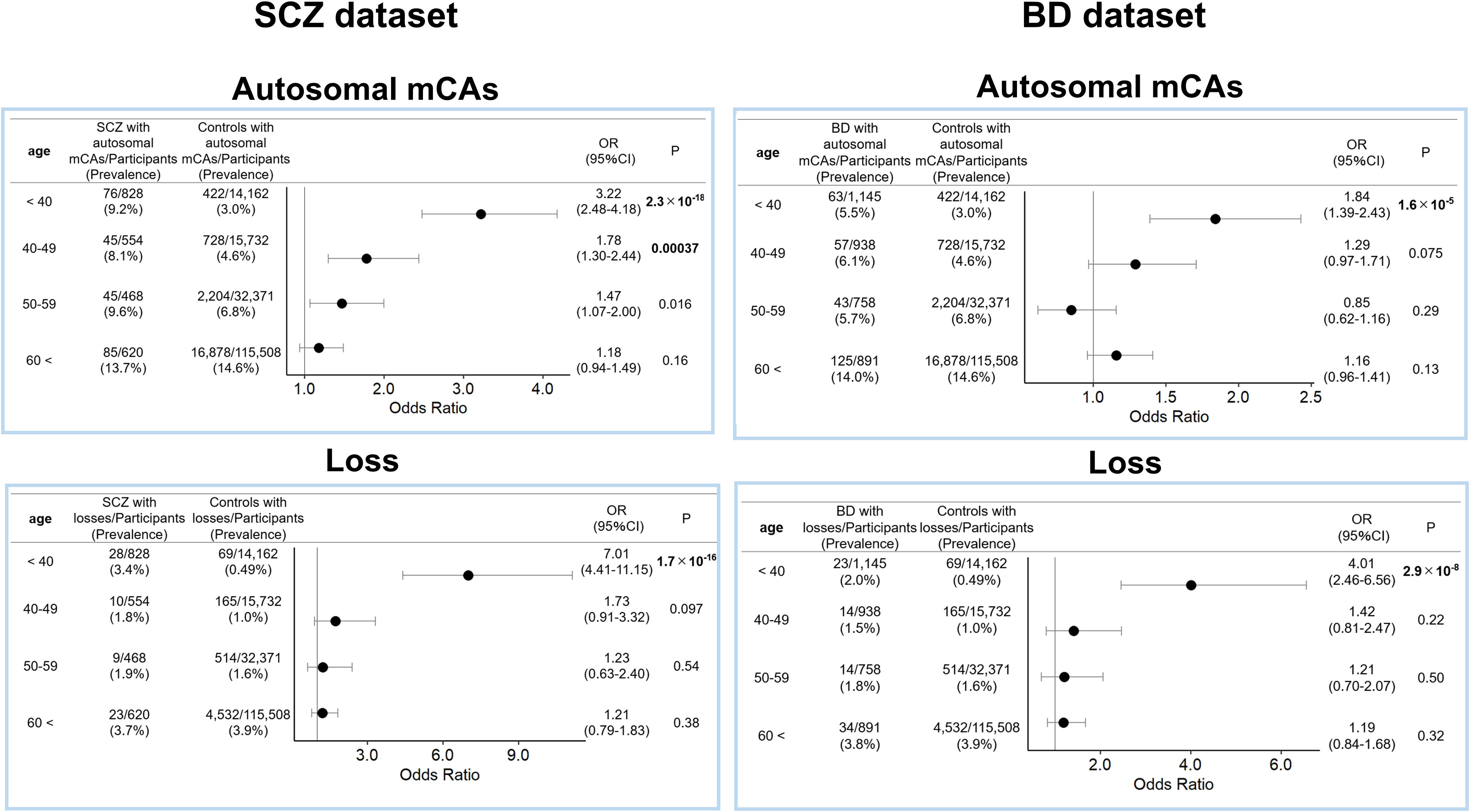
Associations between autosomal mCAs or mosaic loss events and SCZ or BD stratified in each age subgroup. Associations between autosomal mCAs or mosaic loss events and SCZ or BD were evaluated using the subjects in each age category referring to controls. Odds ratio are indicated by dots, and 95%CIs are indicated by two-sided lines. mCAs, mosaic chromosomal alterations; SCZ, schizophrenia; BD, bipolar disorder.

### Significant positive associations of early onset SCZ or BD with mosaic loss events

Based on the findings above, we hypothesized that SCZ with early onset was positively associated with autosomal mCAs or mosaic losses compared with SCZ with late onset. To test this hypothesis, we extracted patients whose information of age at onset was available and analyzed the associations of mCAs between the SCZ subtypes (SCZ of early or late onset) and each mosaic type (METHODS). As expected, autosomal mCAs, especially mosaic losses, were positively associated with SCZ of early onset with higher effect sizes than those for SCZ without stratification. Furthermore, autosomal mCAs were rather negatively associated with late onset SCZ (Supplementary Table S2). These associations were consistent in the BD dataset. These results indicate that the contributions of mCAs differ between early and late onset of these psychiatric diseases.

### Significant and CF-dependent associations of autosomal mCAs or mosaic losses with SCZ or BD

We next analyzed cell fraction (CF) of autosomal mCAs, reflecting clone sizes of expanded cell with mutations. As a result, we figured out both diseases displayed higher CF of autosomal mCAs than controls. Especially, CF of mosaic losses drove the differences in SCZ (Supplementary Table S3 and Fig. S3). Since high CF of mCA would result in strong biological influence of mCA, we hypothesized that the effect sizes of mCAs on SCZ increased in a CF-dependent manner. We extracted subjects with SCZ whose autosomal CF > 5% or > 10%. The effect sizes of autosomal mCA with CF > 5% or 10% were higher than those without the CF threshold, indicating that the association between autosomal mCA and SCZ increased in a CF-dependent manner (Fig. 4). The significant CF-dependent association with SCZ was also observed in mosaic loss events (Fig. 4). We confirmed that these associations were consistent when we analyzed the BD dataset (Fig. 4). When further dissecting this analysis by early or late onset disease phenotypes (METHODS), we found early onset SCZ or BD primarily drove the CF-dependent increased burden (Supplementary Fig. S4 and S5).

**Figure 4.**
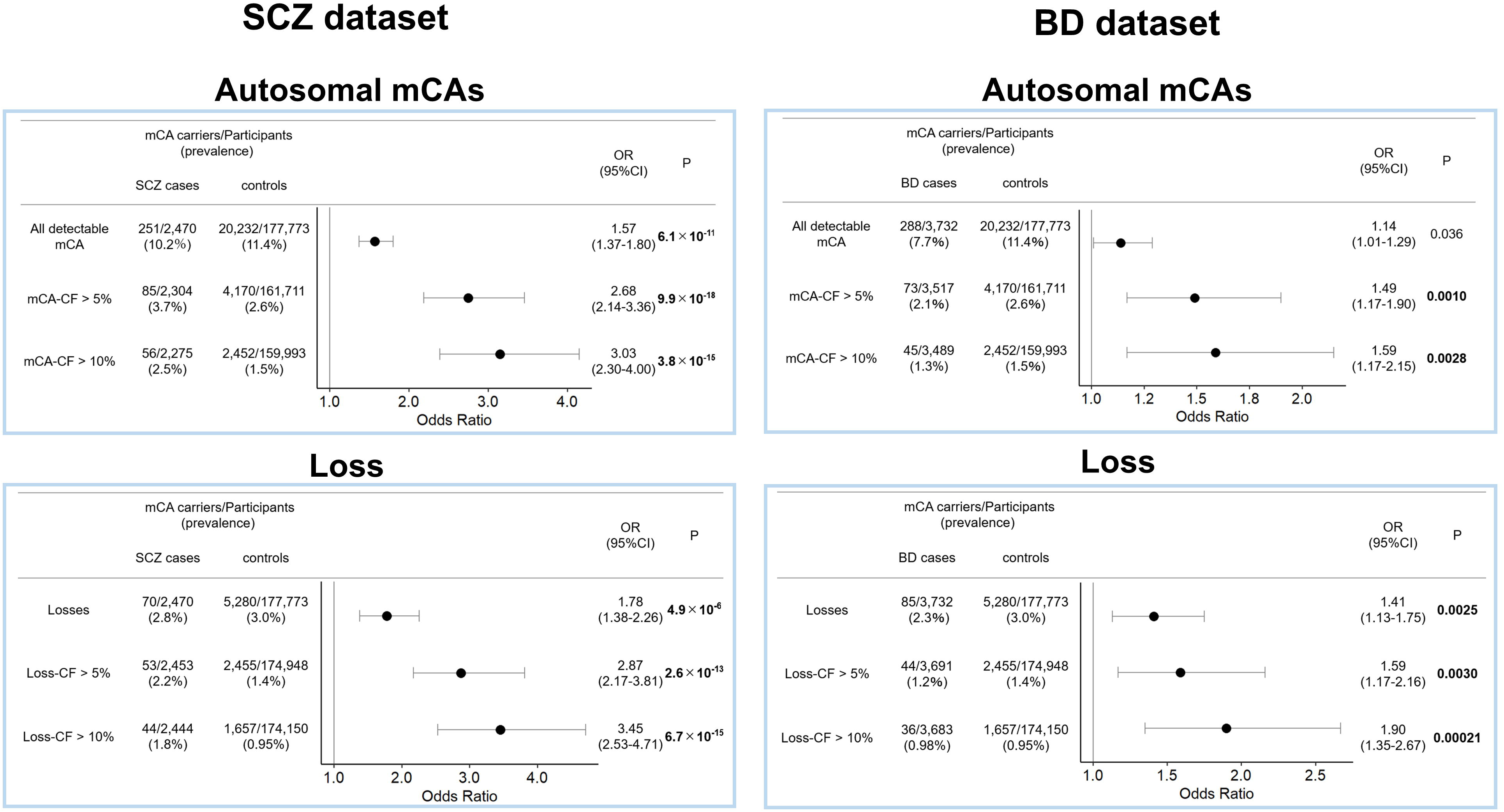
Associations between autosomal mCAs or mosaic loss events with CF-threshold and SCZ or BD. Associations between autosomal mCAs-CF > 5% or > 10% and SCZ or BD were evaluated referring to controls (upper left and upper right). Associations between loss-CF > 5% or > 10% and SCZ or BD were also evaluated referring to controls (lower left and lower right). The associations between autosomal mCAs or losses without CF threshold and SCZ or BD were also shown. Odds ratio are indicated by dots, and 95%CIs are indicated by two-sided lines. mCAs, mosaic chromosomal alterations; SCZ, schizophrenia; BD, bipolar disorder.

### Mosaic losses with high CF located the cytoband 4q35.2 region in SCZ

To evaluate the gene involvement affected by the mosaic loss events, we performed the CMH test using hg38 gene list as a reference (METHODS). We found statistically significant associations for the mosaic loss events in cytoband 4q35.2, where the notable affected genes are *DUX4L, FRG1,* and *FRG2* (P=4.12×10^-8^) (Supplementary Table S4). The events were observed in 10 SCZ cases. Of 10 cases, 7 cases have mosaic losses with CF > 10% ranging from 11.9% to 31.5%, suggesting substantial contribution of the mCA in this region to SCZ. The effect sizes of mosaic losses in cytoband 4q35.2 with SCZ of early onset were higher than those for SCZ without stratification (Supplementary Table S5). On the other hand, we did not identify a significant association in patients with BD.

### Significant positive associations of autosomal mCAs and mosaic losses with SCZ or BD conditioning on genetic risks

To evaluate the associations between autosomal mCAs or losses and SCZ or BD conditioning on the genetic risks, we calculated the genetic risk scores for SCZ and BD (METHODS). We found no significant associations between genetic risk scores and autosomal mCA or mosaic loss events in each dataset (Supplementary Table S6). The logistic regression model revealed autosomal mCAs and mosaic loss events were significantly associated with SCZ conditioning on the genetic risk scores (Supplementary Table S7). These association patterns were consistent in BD (Supplementary Table S7).

### CF-dependent and significant antagonistic interactive association of autosomal mCA or mosaic loss events and PRS on SCZ

To evaluate the interactions between autosomal mCAs and polygenic risk scores (PRS) with SCZ, we stratified the patients into six subgroups based on the PRS status and mCA status. Then, we calculated the risk of SCZ in each subgroup referring to low PRS and mCA negative subgroup using logistic regression models adjusted for sex and age. While the SCZ risk significantly increased in an autosomal CF-dependent manner in the low PRS subgroup, the risk just steadily increased in the high PRS subgroup (Fig. 5). This association pattern was also observed in autosomal mosaic loss events (Fig. 5). Statistically, we evaluated the interaction between autosomal mCAs or losses and PRS with SCZ using one-way ANOVA (METHODS). We found a statistically significant interaction between autosomal mCA-CF or loss-CF >5% and PRS in the SCZ (autosomal mCAs; P=0.0010, losses; P=0.0098), (Supplementary Table S8). These interactions were regarded as antagonistic interactions since the excess risk ratio among individuals with both factors (PRS group high and mCA-CF>5%) is less than the sum of the excess risk ratio of each factor considered alone. There was no statistically significant interaction in the BD (Supplementary Table S8 and Fig. S6).

**Figure 5.**
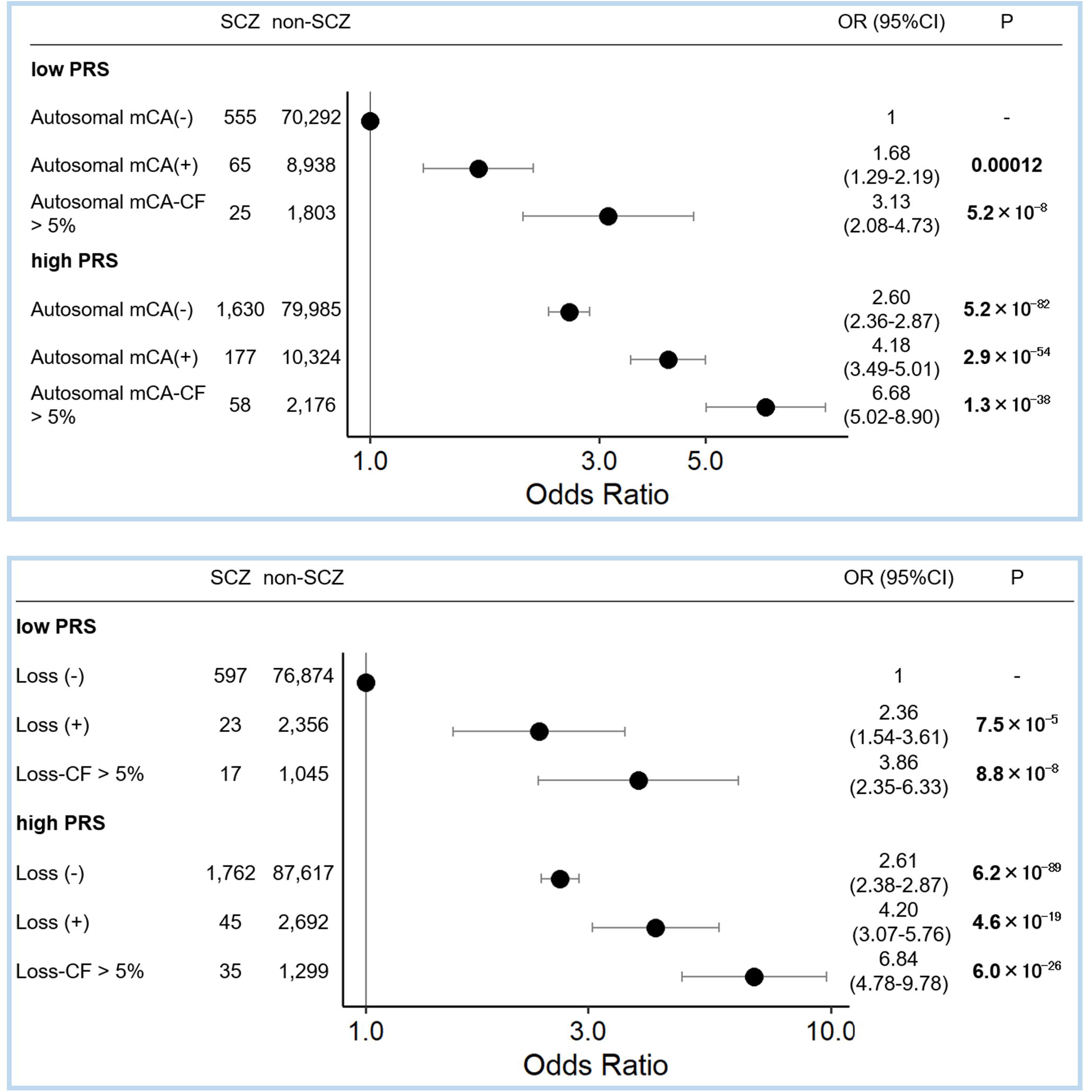
Combinatory associations of PRS and autosomal mCAs or mosaic loss events on SCZ. The associations of the PRS subgroups with SCZ stratified by mCA status were evaluated referring to low PRS subgroup without mCA. Autosomal mCAs (upper) or mosaic loss events (lower) were separately assessed. Odds ratio are indicated by dots, and 95%CIs are indicated by two-sided lines. mCA, mosaic chromosomal alteration, SCZ, schizophrenia.

## DISCUSSION

The associations between mCAs in peripheral blood and SCZ have been explored in previous studies [9, 10]. Consistent with the previous studies, we found the positive associations between mCAs, particularly mosaic loss events, and SCZ. However, the previous studies lacked the information of age and could not definitively compare the frequency of mCAs between cases and controls by conditioning on age. Additionally, the associations between mCAs and BD have never been explored. In the present study, we found that autosomal mCAs, particularly mosaic losses, are positively associated with SCZ or BD of young onset. We also found the possible interaction between mosaic losses and PRS for SCZ.

To reveal the potential risk of mCAs in psychiatric disorders, brain tissue is the best option to study. However, due to the difficulties of the sample collection, we often alternatively use blood samples. Previous studies have found that high CF of mosaic events might reflect the mCAs in the early developmental period [28], and these mosaic events are shared across the tissues [12] (from the brain to the peripheral blood). In the present study, we detected mCAs from peripheral blood and found that mosaic losses were positively associated with SCZ. These associations were supported by the findings in mCA with high fraction. In addition, the positive associations between mosaic losses and SCZ were highlighted in the groups of young age and young onset. These common results across the ancestries, namely, associations between SCZ and autosomal mosaic loss in peripheral blood, suggest that the early development of mosaic losses play a pivotal role for the pathogenesis of SCZ. Considering decreasing differences in mCA positivity between SCZ and controls in aged subgroups, mCA in SCZ at young onset may share the same basics of development as age-related mCA. SCZ of early onset were characterized by higher severity of positive symptoms or worse treatment response compared with SCZ of late onset [17]. These differences in clinical phenotypes would partially be explained by the difference in mosaic burdens between them. Furthermore, the elevated mosaic loss burden in BD suggests a common somatic pathophysiological mechanism between these psychiatric diseases in addition to the shared genetic architecture.

It is well known that genetic and environmental factors and their interactions are associated with the development of SCZ [1]. Regarding the environmental factors, obstetric complications, infections, or seasons at birth are reported to be associated with SCZ. The observed significant possible interaction between the high burden of mosaic events and genetic risks for SCZ would support the idea that mosaic losses are one of the environmental (somatic) risk factors for SCZ based on known interactions between environmental and genetic components in SCZ development. Interestingly, our previous study showed similar interactions in rheumatoid arthritis, suggesting potential generalizability of this interaction in diverse phenotypes [20].

Regarding the biological mechanisms of the mCAs in psychiatric diseases, Sherman et al. [29] detected mCAs from peripheral blood of probands with autism spectrum disorder (ASD) and unaffected siblings and evaluated the contributions of mCAs to the risks of ASD. They found that the patients with ASD have a larger size of mCAs compared with unaffected siblings and also found a positive correlation between the size of mCAs and the disease severity [29]. Another study detected mosaic mutations in the brains of patients with ASD, and they found ASD has excess mosaic mutations in the enhancer regions associated with genes that have brain-specific expressions [30]. Similarly, Maury et al. [11] have found that somatic mutations in the open chromatin region increased in the brain of SCZ, and some of these mutations alter the gene expression, including SCZ risk genes. These previous findings suggest how mCAs potentially contribute to development of psychiatric diseases. In the present study, we identified that *DUX4L, FRG1,* and *FRG2* in 4q35.2 were associated with mosaic losses in patients with SCZ. However, these genes were reported as candidate genes of facioscapulohumeral muscular dystrophy [31] and we cannot conclude the direct pathophysiological mechanism of these genes to SCZ.

Additionally, an increase in mCAs in these psychiatric diseases would be associated with abnormal aging since mCAs accumulate with age [19]. Consistent with this hypothesis, previous reports have found associations between SCZ and the shortened telomere length [32] and the epigenetic clock. Future studies to evaluate biological aging (telomere length or epigenetic clock) will clarify the mechanism of mCAs with SCZ.

Our study had several limitations. First, we did not replicate the associations between mCAs and SCZ or BD using other independent datasets. However, the consistent results with the previous studies support our findings. Second, we did not directly detect mCAs in the brain because human brain tissue is difficult to obtain. The present study using blood samples could not identify the direct reasonable causal mechanisms between mCAs and SCZ risk genes; future studies using brain tissues would allow further characterization of the potential risk of mCAs in SCZ or BD.

To the best of our knowledge, this is the first study to demonstrate the common somatic architecture between SCZ and BD particularly in onset of young age, as well as the possible interactions between mosaic losses and SCZ genetic components, which may explain the contribution of genetic and somatic architecture in patients with SCZ.

## Acknowledgements

This work was supported by the Japan Agency for Medical Research and Development (AMED) grants 21ek0109555 (C.T.), 21tm0424220 (C.T.), 21ck0106642 (C.T.), 23ek0410114 (C.T.), 23tm0424225 (C.T.), JP21wm0425008 (N.I. and M.I.), JP23tm0524001 (M.I.), JP21wm0525024 (M.I.), 21tm0424220 (M.I.) and JP23dk0307123 (M.I.); the Japan Society for the Promotion of Science (JSPS) KAKENHI grant JP20H00462 (C.T.), JP21H02854 (M.I.), 24K02381 (M.I.) and JP22H03003 (N.I.); and Takeda Hosho Grants for Research in Medicine. The cartoons shown in Fig. 1 were created using Bio-Render.com. We thank the staff of BBJ for collecting and managing the samples and clinical information.

## Conflict of Interest

S.U., T.S., X.L., Y.I., K.H., M.I., N.I. and C.T. have no conflicts of interest. G.G. declared competing interests, and patent application PCT/WO2019/079493 has been filed for the mosaic chromosomal alteration detection method used in this study.

## Data availability

Statistical codes are available from Chikashi Terao (ORCID 0000-0002-6452-4095) at any time, only on reasonable request. The Mosaic Chromosomal Alterations (MoChA) pipelines used for mosaic calling (mocha.wdl) are available at (https://github.com/freeseek/mochawdl). The genotype and IDAT data of BBJ used for this research was available at the website of the NBDC Human Database (https://humandbs.dbcls.jp/en/) of the Database Center for Life Science (DBCLS) / the Joint Support-Center for Data Science Research (DS) of the Research Organization of Information and Systems (ROIS). The dataset ID are JGAD000836 and JGAS000114.

## Author contributions

M.I., N.I. and C.T. conceived the project. S.U. and C.T. analysed the data. G.G. developed MoChA pipelines for mosaic calling. T.S collected the detailed clinical information. S.U., T.S., X.L., Y.I., K.H., and C.T. wrote the manuscript. All authors have critically reviewed and approved the final version of the manuscript.

## Ethics approval

All patients provided informed consent, and this study was approved by the IRB of RIKEN Center for Integrative Medical Sciences.

## Notes

### Author Declarations

IRB of RIKEN Center for Integrative Medical Sciences gave ethical approval for this work

## REFERENCES

1. Robinson N, Bergen SE. Environmental Risk Factors for Schizophrenia and Bipolar Disorder and Their Relationship to Genetic Risk: Current Knowledge and Future Directions. Front Genet 2021; 12: 686666.

2. Trubetskoy V, Pardinas AF, Qi T, Panagiotaropoulou G, Awasthi S, Bigdeli TB et al. Mapping genomic loci implicates genes and synaptic biology in schizophrenia. Nature 2022; 604(7906): 502–508.

3. Vijg J, Dong X. Pathogenic Mechanisms of Somatic Mutation and Genome Mosaicism in Aging. Cell 2020; 182(1): 12–23.

4. Bizzotto S, Walsh CA. Genetic mosaicism in the human brain: from lineage tracing to neuropsychiatric disorders. Nat Rev Neurosci 2022; 23(5): 275–286.

5. Bae T, Tomasini L, Mariani J, Zhou B, Roychowdhury T, Franjic D et al. Different mutational rates and mechanisms in human cells at pregastrulation and neurogenesis. Science 2018; 359(6375): 550–555.

6. Lodato MA, Rodin RE, Bohrson CL, Coulter ME, Barton AR, Kwon M et al. Aging and neurodegeneration are associated with increased mutations in single human neurons. Science 2018; 359(6375): 555–559.

7. D’Gama AM, Walsh CA. Somatic mosaicism and neurodevelopmental disease. Nat Neurosci 2018; 21(11): 1504–1514.

8. Liu X, Kamatani Y, Terao C. Genetics of autosomal mosaic chromosomal alteration (mCA). J Hum Genet 2021; 66(9): 879–885.

9. Maury EA, Sherman MA, Genovese G, Gilgenast TG, Kamath T, Burris SJ et al. Schizophrenia-associated somatic copy-number variants from 12,834 cases reveal recurrent NRXN1 and ABCB11 disruptions. Cell Genom 2023; 3(8): 100356.

10. Chang K, Jian X, Wu C, Gao C, Li Y, Chen J et al. The Contribution of Mosaic Chromosomal Alterations to Schizophrenia. Biol Psychiatry 2024.

11. Maury EA, Jones A, Seplyarskiy V, Nguyen TTL, Rosenbluh C, Bae T et al. Somatic mosaicism in schizophrenia brains reveals prenatal mutational processes. Science 2024; 386(6718): 217–224.

12. Ju YS, Martincorena I, Gerstung M, Petljak M, Alexandrov LB, Rahbari R et al. Somatic mutations reveal asymmetric cellular dynamics in the early human embryo. Nature 2017; 543(7647): 714–718.

13. Ikeda M, Aleksic B, Kirov G, Kinoshita Y, Yamanouchi Y, Kitajima T et al. Copy number variation in schizophrenia in the Japanese population. Biol Psychiatry 2010; 67(3): 283–286.

14. Ikeda M, Aleksic B, Kinoshita Y, Okochi T, Kawashima K, Kushima I et al. Genome-wide association study of schizophrenia in a Japanese population. Biol Psychiatry 2011; 69(5): 472–478.

15. Ikeda M, Takahashi A, Kamatani Y, Okahisa Y, Kunugi H, Mori N et al. A genome-wide association study identifies two novel susceptibility loci and trans population polygenicity associated with bipolar disorder. Mol Psychiatry 2018; 23(3): 639–647.

16. Nagai A, Hirata M, Kamatani Y, Muto K, Matsuda K, Kiyohara Y et al. Overview of the BioBank Japan Project: Study design and profile. J Epidemiol 2017; 27(3S): S2–S8.

17. Maglione JE, Thomas SE, Jeste DV. Late-onset schizophrenia: do recent studies support categorizing LOS as a subtype of schizophrenia? Curr Opin Psychiatry 2014; 27(3): 173–178.

18. Arnold I, Dehning J, Grunze A, Hausmann A. Old Age Bipolar Disorder-Epidemiology, Aetiology and Treatment. Medicina (Kaunas) 2021; 57(6).

19. Terao C, Suzuki A, Momozawa Y, Akiyama M, Ishigaki K, Yamamoto K et al. Chromosomal alterations among age-related haematopoietic clones in Japan. Nature 2020; 584(7819): 130–135.

20. Uchiyama S, Ishikawa Y, Ikari K, Honda S, Hikino K, Tanaka E et al. Mosaic loss of chromosome Y characterises late-onset rheumatoid arthritis and contrasting associations of polygenic risk score based on age at onset. Ann Rheum Dis 2025.

21. Genomes Project C, Auton A, Brooks LD, Durbin RM, Garrison EP, Kang HM et al. A global reference for human genetic variation. Nature 2015; 526(7571): 68–74.

22. Ito S, Liu X, Ishikawa Y, Conti DD, Otomo N, Kote-Jarai Z et al. Androgen receptor binding sites enabling genetic prediction of mortality due to prostate cancer in cancer-free subjects. Nat Commun 2023; 14(1): 4863.

23. Purcell S, Neale B, Todd-Brown K, Thomas L, Ferreira MA, Bender D et al. PLINK: a tool set for whole-genome association and population-based linkage analyses. Am J Hum Genet 2007; 81(3): 559–575.

24. Delaneau O, Zagury JF, Robinson MR, Marchini JL, Dermitzakis ET. Accurate, scalable and integrative haplotype estimation. Nat Commun 2019; 10(1): 5436.

25. Ge T, Chen CY, Ni Y, Feng YA, Smoller JW. Polygenic prediction via Bayesian regression and continuous shrinkage priors. Nat Commun 2019; 10(1): 1776.

26. Lam M, Chen CY, Li Z, Martin AR, Bryois J, Ma X et al. Comparative genetic architectures of schizophrenia in East Asian and European populations. Nat Genet 2019; 51(12): 1670–1678.

27. Mullins N, Forstner AJ, O’Connell KS, Coombes B, Coleman JRI, Qiao Z et al. Genome-wide association study of more than 40,000 bipolar disorder cases provides new insights into the underlying biology. Nat Genet 2021; 53(6): 817–829.

28. Lodato MA, Woodworth MB, Lee S, Evrony GD, Mehta BK, Karger A et al. Somatic mutation in single human neurons tracks developmental and transcriptional history. Science 2015; 350(6256): 94–98.

29. Sherman MA, Rodin RE, Genovese G, Dias C, Barton AR, Mukamel RE et al. Large mosaic copy number variations confer autism risk. Nat Neurosci 2021; 24(2): 197–203.

30. Rodin RE, Dou Y, Kwon M, Sherman MA, D’Gama AM, Doan RN et al. The landscape of somatic mutation in cerebral cortex of autistic and neurotypical individuals revealed by ultra-deep whole-genome sequencing. Nat Neurosci 2021; 24(2): 176–185.

31. Hansda AK, Tiwari A, Dixit M. Current status and future prospect of FSHD region gene 1. J Biosci 2017; 42(2): 345–353.

32. Russo P, Prinzi G, Proietti S, Lamonaca P, Frustaci A, Boccia S et al. Shorter telomere length in schizophrenia: Evidence from a real-world population and meta-analysis of most recent literature. Schizophr Res 2018; 202: 37–45.

