## Supplementary information for "Associations between mosaic loss and schizophrenia or bipolar disorder of young onset"

**Supplementary Figures**

**S1.** Associations between CN-LOHs, gains, mLOX, or mLOY and SCZ stratified by age

**S2.** Associations between CN-LOHs, gains, mLOX, or mLOY and BD stratified by age

**S3.** Box plot of cell fraction of mosaic events in SCZ and BD dataset

**S4.** Associations between autosomal mCAs or mosaic loss events with CF-threshold and early onset SCZ or BD

**S5.** Associations between autosomal mCAs or mosaic loss events with CF-threshold and late onset SCZ or BD

**S6.** Combinatory associations of GRS and autosomal mCAs or mosaic loss events on BD

**Supplementary Tables**

**S1.** Demographic features of participants in each dataset

**S2.** Associations between different detectable mCAs and SCZ or BD based on the onset age of the disease

**S3.** The cell fraction of cases and controls in each dataset

**S4**. Associations between mosaic losses and genes on the specific locus in SCZ

**S5**. Associations of mosaic losses in cytoband 4_q35.2 with early onset SCZ

**S6**. Association between autosomal mCA or mosaic loss events and genetic risk in each dataset

**S7.** Associations of autosomal mCA or mosaic loss events and SCZ-PRS or BD-GRS with disease subsets

**S8.** Results of the one-way ANOVA to evaluate the goodness of fit between the two models

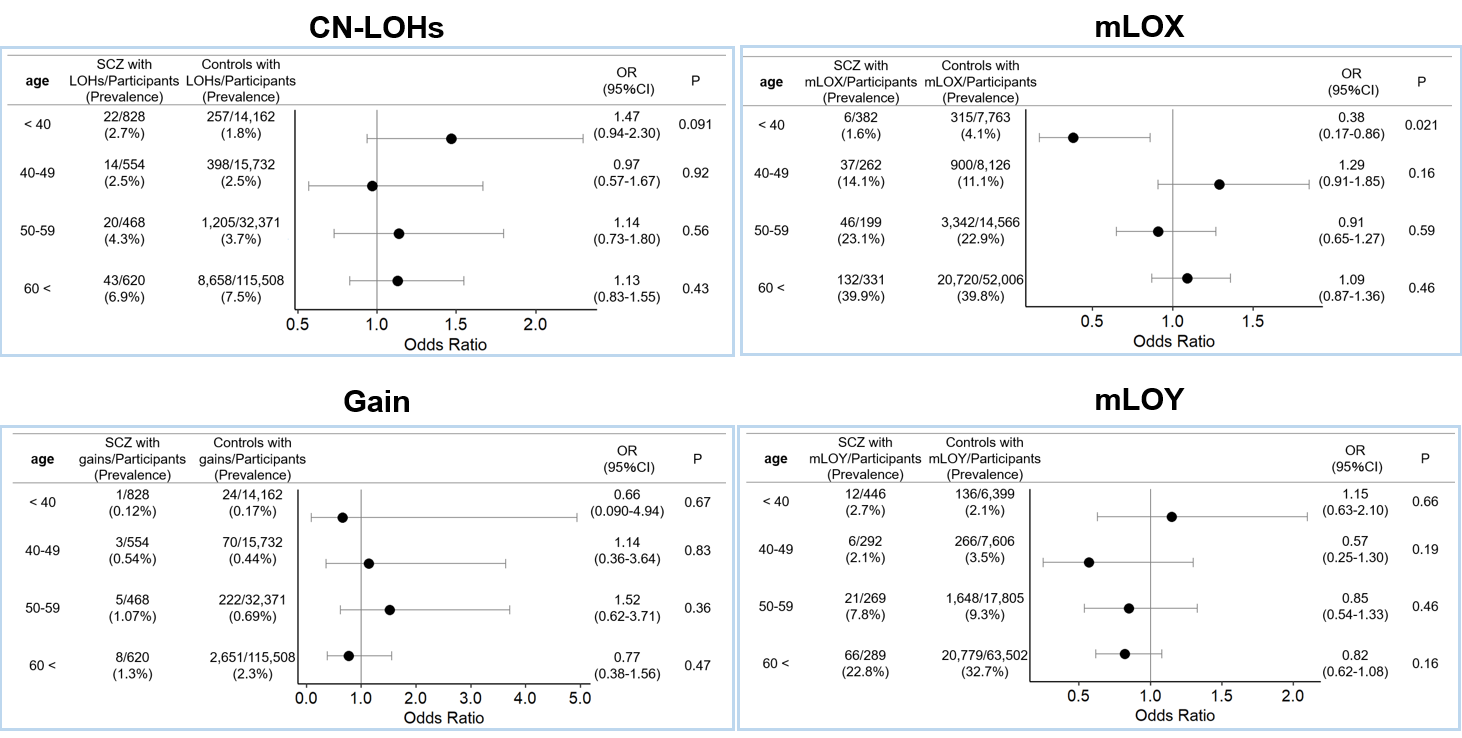
**Supplementary Figure S1. Associations between CN-LOHs, gains, mLOX, or mLOY and SCZ stratified by age.**

Associations between CN-LOHs, gains, mLOX, or mLOY and SCZ were evaluated in each age subgroup referring to controls. ORs were indicated by dots and 95%CIs were indicated by two-sided lines. SCZ, schizophrenia; CN-LOHs, copy-neutral loss of heterozygosity; mLOX, mosaic loss of chromosome X; mLOY, mosaic loss of chromosome Y.

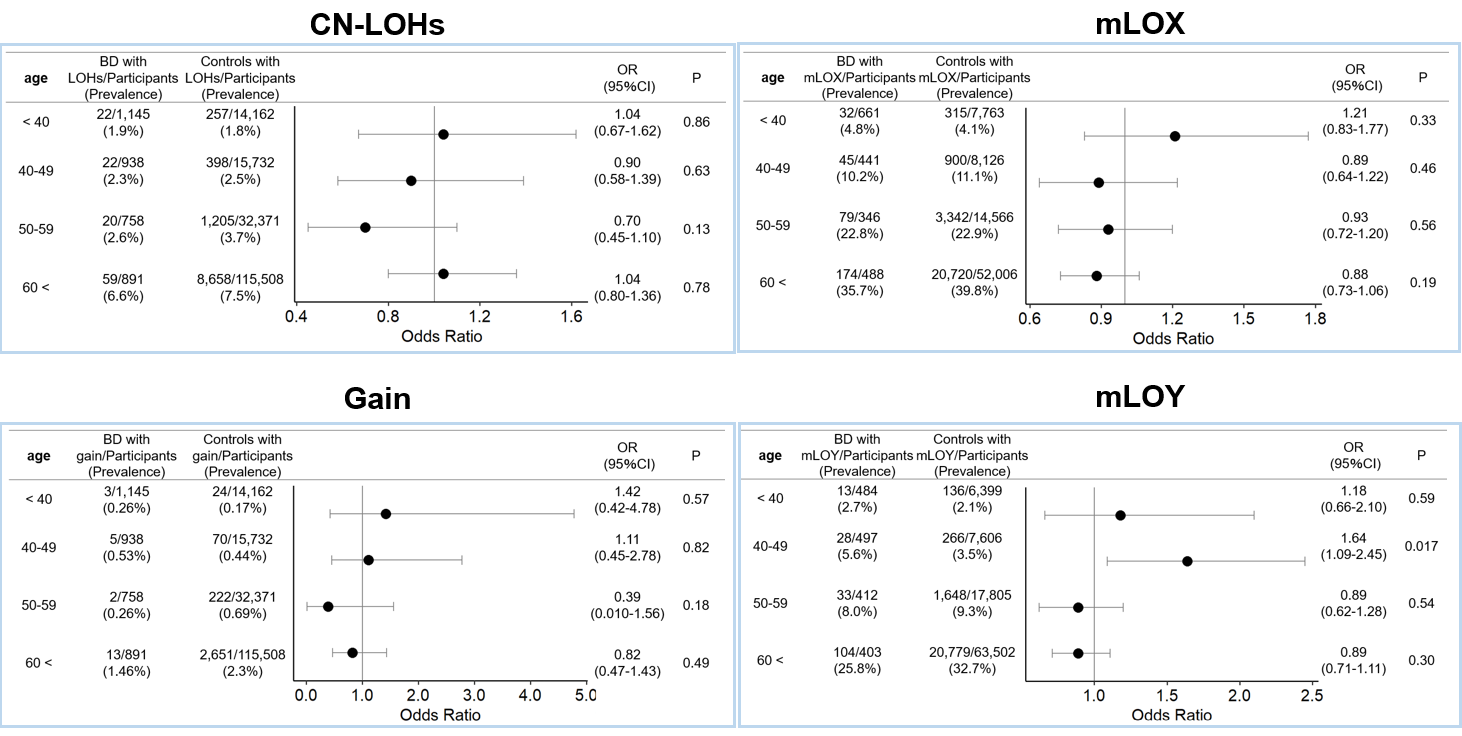
**Supplementary Figure S2. Associations between CN-LOHs, gains, mLOX, or mLOY and BD stratified by age.**

Associations between CN-LOHs, gains, mLOX, or mLOY and BD were evaluated in each age subgroup referring to controls. ORs were indicated by dots and 95%CIs were indicated by two-sided lines. BD, bipolar disorders; CN-LOHs, copy-neutral loss of heterozygosity; mLOX, mosaic loss of chromosome X; mLOY, mosaic loss of chromosome Y.

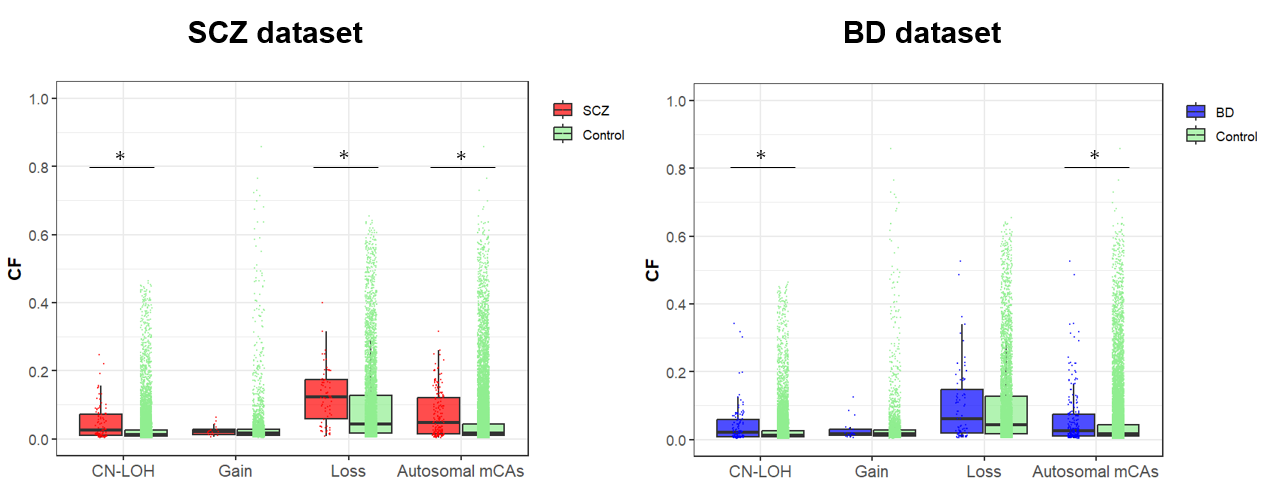
**Supplementary Figure S3. Box plot of cell fraction of mosaic events in SCZ and BD dataset.**

Box plot of cell fraction of mosaic events in cases vs. controls among SCZ and BD dataset were shown.

* P < 0.05, Wilcoxon rank sum test. SCZ, schizophrenia; BD, bipolar disorders; CF, cell fraction; CN-LOH, copy-neutral loss of heterozygosity; **mCAs, mosaic chromosomal alterations.**

**Supplementary Figure S4. Associations between autosomal mCAs or mosaic loss events with CF-threshold and early onset SCZ or BD.**

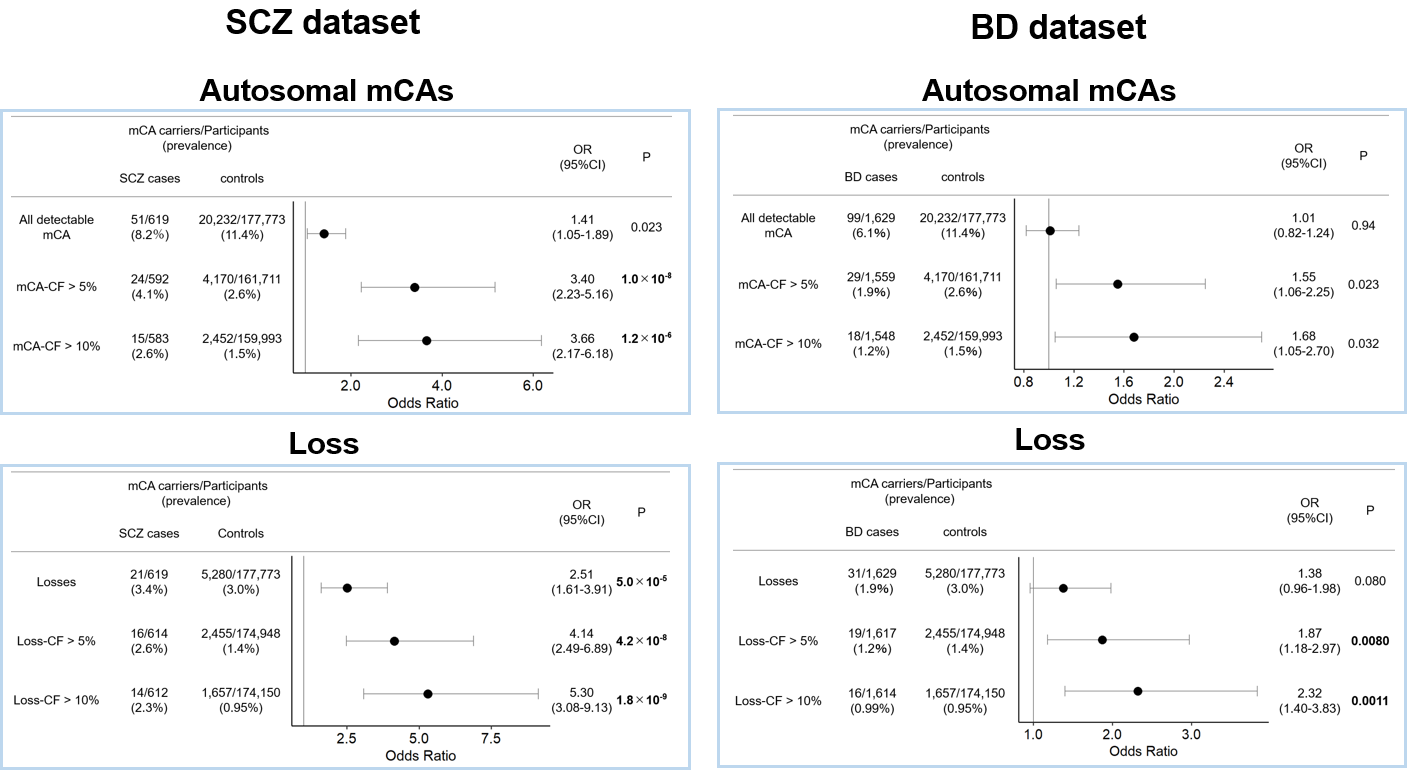

Associations between autosomal mCAs-CF > 5% or > 10% and early onset SCZ or BD were evaluated referring to controls (upper left and upper right). Associations between loss-CF > 5% or > 10% and early onset SCZ or BD were also evaluated referring to controls (lower left and lower right). The associations between autosomal mCAs or losses without CF threshold and early onest SCZ or BD were also shown. Odds ratio are indicated by dots, and 95%CIs are indicated by two-sided lines. mCAs, mosaic chromosomal alterations; SCZ, schizophrenia; BD, bipolar disorder.

**Supplementary Figure S5. Associations between autosomal mCAs or mosaic loss events with CF-threshold and late onset SCZ or BD**.

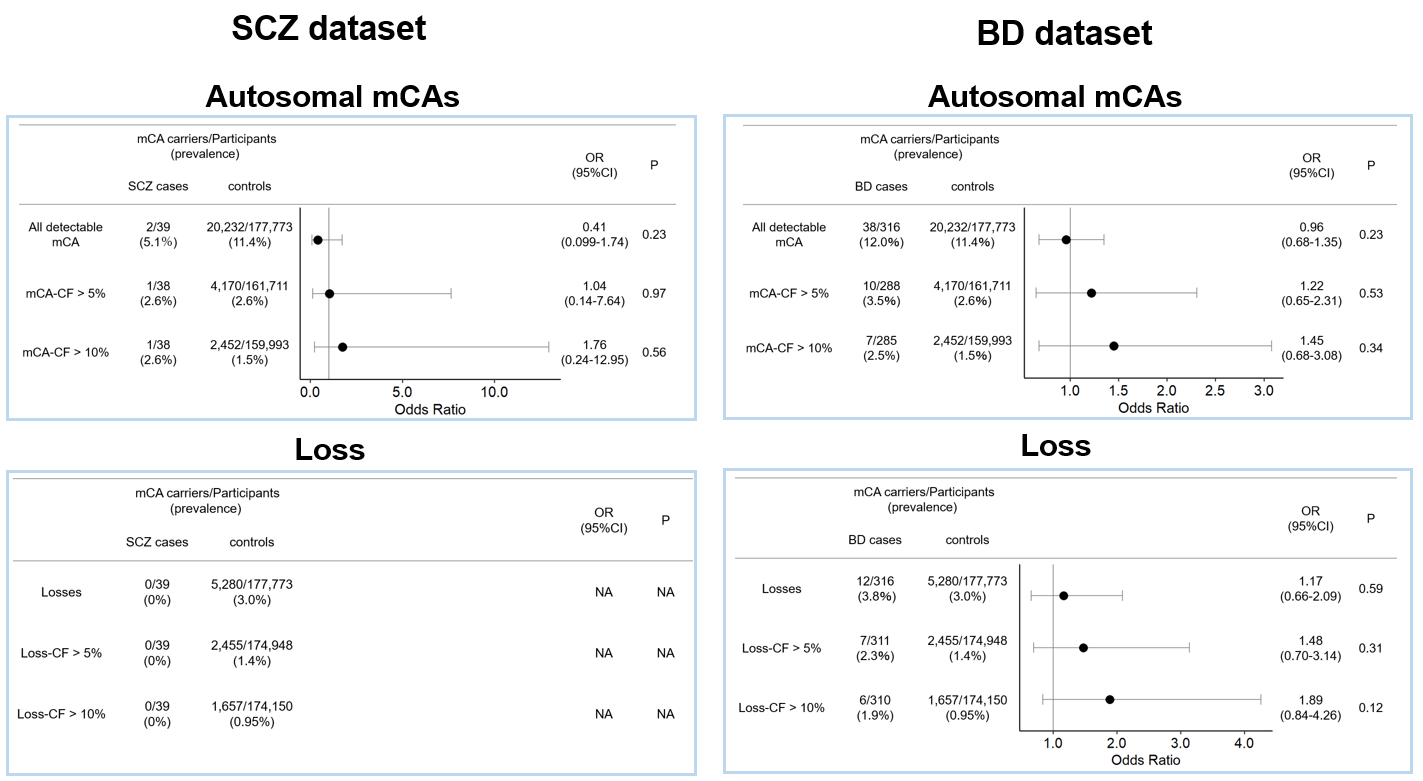

Associations between autosomal mCAs-CF > 5% or > 10% and late onset SCZ or BD were evaluated referring to controls (upper left and upper right). Associations between loss-CF > 5% or > 10% and late onset SCZ or BD were also evaluated referring to controls (lower left and lower right). The associations between autosomal mCAs or losses without CF threshold and late onest SCZ or BD were also shown. Odds ratio are indicated by dots, and 95%CIs are indicated by two-sided lines. mCAs, mosaic chromosomal alterations; SCZ, schizophrenia; BD, bipolar disorder.

**Supplementary Figure S6. Combinatory associations of GRS and autosomal mCAs or mosaic loss events on BD.**

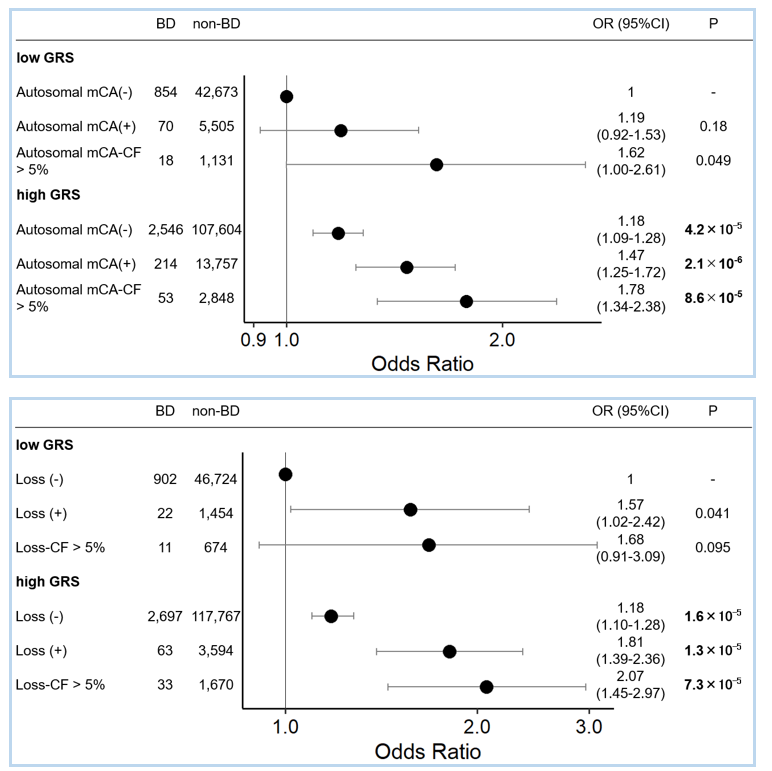

The associations of the GRS subgroups with BD stratified by mCA status were evaluated referring to low GRS subgroup without mCA. Autosomal mCAs (upper) or mosaic loss events (lower) were separately assessed. Odds ratio are indicated by dots, and 95%CIs are indicated by two-sided lines. mCA, mosaic chromosomal alteration, BD, bipolar disorders.

**Supplementary Table S1.** **Demographic features of participants in each dataset.**

|  | **SCZ dataset** | | **BD dataset** | |
| --- | --- | --- | --- | --- |
|  | **SCZ** | **Controls** | **BD** | **Controls** |
| Number of subjects | 2,470 | 177,773 | 3,732 | 177,773 |
| Age (years), median (IQR) | 47.0 [36.0-60.0] | 65.0 [55.0-73.0] | 60.0 [51.0-67.0] | 65.0 [55.0-73.0] |
| Sex, n (%) |  |  |  |  |
| Male | 1,296 (52.5%) | 95,312 (53.6%) | 1,796 (48.1%) | 95,312 (53.6%) |
| Female | 1,174 (47.5%) | 82,461 (46.4%) | 1,936 (51.9%) | 82,461 (46.4%) |
| Types of diseases, n (%) |  |  |  |  |
| Early onset | 619 (25.1%) | NA | 1,629 (43.6%) | NA |
| Late onset | 39 (1.5%) | NA | 316 (8.5%) | NA |
| No information of age at disease onset | 1,812 (73.4%) | NA | 1,787 (47.9%) | NA |
| Types of arrays, n (%) |  |  |  |  |
| OEE v1.0 | 2,052 (83.1%) | 34,426 (19.4%) | 1,459 (39.1%) | 34,426 (19.4%) |
| OEE v1.2 | 418 (16.9%) | 111,209 (62.6%) | 1,478 (39.6%) | 111,209 (62.6%) |
| OEE v1.4 | NA | NA | 562 (15.1%) | NA |
| OEE v1.6 | NA | NA | 233 (6.2%) | NA |
| OE v1.0 and HE v1.0, 1.1 | NA | 32,138 (18.0%) | NA | 32,138 (18.0%) |

**SCZ, schizophrenia; BD, bipolar disorder; OEE, OmniExpress Exome; OE, OmniExpress; HE, HumanExome.**

**Supplementary Table S2. Associations between different detectable mCAs and SCZ or BD based on the onset age of the disease.**

|  |  | mCA carriers/Partcipants (Prevalence) | | OR  (95%CI) | P | mCA carriers/Partcipants (Prevalence) | | OR  (95%CI) | P |
| --- | --- | --- | --- | --- | --- | --- | --- | --- | --- |
|  |  | SCZ cases | non-SCZ controls |  |  | BD cases | non-BD controls |  |  |
| Autosomal  mCAs | early-onset | 51/619  (8.2%) | 20,232/177,773  (11.4％) | 1.41  (1.05-1.89) | 0.023 | 99/1,629  (6.1%) | 20,232/177,773 (11.4％) | 1.01  (0.82-1.24) | 0.94 |
|  | late-onset | 2/39  (5.1%) |  | 0.41  (0.099-1.74) | 0.23 | 38/316  (12.0%) |  | 0.96  (0.68-1.35) | 0.23 |
| CN-LOH | early-onset | 19/619  (3.1%) | 10,518/177,773  (5.9％) | 0.93  (0.59-1.48) | 0.76 | 46/1,629  (2.8%) | 10,518/177,773  (5.9％) | 0.86  (0.64-1.15) | 0.76 |
|  | late-onset | 1/39  (2.6%) |  | 0.41  (0.060-3.01) | 0.38 | 15/316  (4.7%) |  | 0.71  (0.42-1.20) | 0.20 |
| Loss | early-onset | 21/619  (3.4%) | 5,280/177,773  (3.0％) | 2.51  (1.61-3.91) | **5.0×10^-5^** | 31/1,629  (1.9%) | 5,280/177,773  (3.0％) | 1.38  (0.96-1.98) | 0.080 |
|  | late-onset | 0/39  (0%) |  | NA | NA | 12/316  (3.8%) |  | 1.17  (0.66-2.09) | 0.59 |
| Gain | early-onset | 4/619  (0.65%) | 2,967/177,773  (1.7％) | 0.95  (0.36-2.57) | 0.93 | 9/1,629  (0.55%) | 2,967/177,773  (1.7％) | 0.82  (0.42-1.59) | 0.56 |
|  | late-onset | 0/39  (0%) |  | NA | NA | 1/316  (0.32%) |  | 0.17  (0.023-1.19) | 0.073 |
| mLOX | early-onset | 42/288  (14.6%) | 25,277/82,461  (30.7%) | 0.89  (0.63-1.27) | 0.53 | 114/833  (13.7%) | 25,277/82,461  (30.7%) | 0.85  (0.69-1.05) | 0.13 |
|  | late-onset | 8/28  (28.6%) |  | 0.79  (0.34-1.85) | 0.59 | 56/182  (30.8%) |  | 0.76  (0.55-1.06) | 0.10 |
| mLOY | early-onset | 21/331  (6.3%) | 22,829/95,312  (24.0%) | 0.66  (0.41-1.04) | 0.074 | 63/796  (7.9%) | 22,829/95,312  (24.0%) | 0.80  (0.61-1.05) | 0.11 |
|  | late-onset | 3/11  (27.3%) |  | 0.93  (0.23-3.75) | 0.92 | 29/134  (21.6%) |  | 0.77  (0.50-1.18) | 0.23 |

OR, odds ratio referring to non-psychiatric disease controls; P value in bold indicate statistical significance based on Bonferroni correction (P<0.008). **SCZ, schizophrenia; BD, bipolar disorder; mCAs, mosaic chromosomal alterations; CN-LOH, copy number-loss of heterozygosity; mLOX, mosaic loss of chromosome X; mLOY, mosaic loss of chromosome Y.**

**Supplementary Table S3. The cell fraction of cases and controls in each dataset corresponding to Supplementary Figure S3.**

|  | SCZ dataset | |  | BD dataset | |  |
| --- | --- | --- | --- | --- | --- | --- |
|  | SCZ | Controls | P | BD | Controls | P |
| CF of CN-LOH, median (IQR) | 0.025  (0.011-0.073) | 0.013  (0.0078-0.027) | **9.9×10^-7^** | 0.021  (0.0091-0.058) | 0.013  (0.0078-0.027) | **0.00013** |
| CF of Gain,  median (IQR) | 0.022  (0.013-0.027) | 0.017  (0.011-0.029) | 0.24 | 0.017  (0.013-0.031) | 0.017  (0.011-0.029) | 0.85 |
| CF of Loss,  median (IQR) | 0.12  (0.059-0.17) | 0.043  (0.018-0.13) | **1.1×10^-6^** | 0.061  (0.019-0.15) | 0.043  (0.018-0.13) | 0.47 |
| CF of  Autosomal mCAs, median (IQR) | 0.048  (0.014-0.12) | 0.017  (0.0098-0.043) | **3.6×10^-12^** | 0.025  (0.010-0.075) | 0.017  (0.0098-0.043) | **0.00066** |

**SCZ, schizophrenia; BD, bipolar disorder;** CF, cell fraction**; CN-LOH, copy number-loss of heterozygosity; mCAs, mosaic chromosomal alterations.** P value in bold indicate statistical significance (P<0.05).

**Supplementary Table S4. Associations between mosaic losses and genes on the specific locus in SCZ**

| Locus | Case  n=2,470 | Control  n=177,773 | OR (95%CI) | P | Notable Genes |
| --- | --- | --- | --- | --- | --- |
| 4_q35.2 | 10 | 94 | 13.2 (5.7-27.5) | **4.1×10^-8^** | *DUX4L, FRG1, FRG2* |

Cases mean the number of patients with SCZ exhibiting the mosaic losses in the specific gene. Controls mean the number of patients with controls exhibiting the mosaic losses in the specific gene. P value in bold indicates statistical significance based on Bonferroni correction (P<1.9×10^-6^).

**Supplementary Table S5. Associations of mosaic losses in cytoband 4_q35.2** **with early onset SCZ**

| Locus | Case  n=619 | Control  n=177,773 | OR (95%CI) | P | Notable Genes |
| --- | --- | --- | --- | --- | --- |
| 4_q35.2 | 3 | 94 | 18.7 (3.5-63.4) | 0.0008 | *DUX4L, FRG1, FRG2* |

Cases mean the number of patients with early onset SCZ exhibiting the mosaic losses in *DUX4L*, *FRG1*, and *FRG2*. Controls mean the number of patients with controls exhibiting the mosaic losses in these specific gene.

**Supplementary Table S6. Association between autosomal mCA or mosaic loss events and genetic risk in each dataset.**

| Dataset | genetic risk score | mosaic | OR (95%CI) | P |
| --- | --- | --- | --- | --- |
| SCZ | PRS | autosomal mCA | 1.02 (0.99-1.04) | 0.14 |
|  |  | Losses | 1.01 (0.97-1.05) | 0.57 |
| BD | GRS | autosomal mCA | 0.97 (0.92-1.02) | 0.23 |
|  |  | Losses | 0.99 (0.89-1.09) | 0.80 |

SCZ, schizophrenia; BD, bipolar disorders, PRS, polygenic risk score; GRS, genetic risk score; mCA, mosaic chromosomal alterations.

**Supplementary Table S7. Associations of autosomal mCA or mosaic loss events and SCZ-PRS or BD-GRS with disease subsets**

|  | Disease type | beta | 95%CI | | P |
| --- | --- | --- | --- | --- | --- |
| PRS | SCZ | 0.84 | 0.78 | 0.89 | **1.2×10^-200^** |
| autosomal mCA |  | 0.44 | 0.30 | 0.58 | **6.3×10^-10^** |
| PRS | SCZ | 0.84 | 0.78 | 0.89 | **4.5×10^-201^** |
| Losses |  | 0.55 | 0.30 | 0.80 | **1.8×10^-5^** |
| GRS | BD | 0.34 | 0.22 | 0.46 | **2.7×10^-8^** |
| autosomal mCA |  | 0.16 | 0.033 | 0.28 | **1.3×10^-2^** |
| GRS | BD | 0.34 | 0.22 | 0.46 | **2.7×10^-8^** |
| Losses |  | 0.37 | 0.15 | 0.59 | **1.1×10^-3^** |

P value in bold indicates statistical significance (P<0.05).

**Supplementary Table S8. Results of the one-way ANOVA to evaluate the goodness of fit between the two models**

| Disease | mosaic type |  | Residual deviance | Deviance | P |
| --- | --- | --- | --- | --- | --- |
| SCZ | autosomal mCA | Model1 | 25041 |  |  |
|  |  | Model2 | 25030 | 10.8 | **0.0010** |
|  | Loss | Model1 | 25045 |  |  |
|  |  | Model2 | 25039 | 6.67 | **0.0098** |
| BD | autosomal mCA | Model1 | 35587 |  |  |
|  |  | Model2 | 35585 | 2.07 | 0.15 |
|  | Loss | Model1 | 35634 |  |  |
|  |  | Model2 | 35633 | 0.64 | 0.43 |

$$model1: SCZ (BD)～ mCA+PRS (GRS) group$$

$$model2: SCZ (BD)～ mCA+PRS \left( GRS \right) group+PRS \left( GRS \right) group\times mCA\mathrm{CF}>5\%$$

The p-value in bold indicates statistical significance (P<0.05).
